# Can we start to ignore the SARS-CoV-2 disease?

**DOI:** 10.1101/2023.09.18.23295709

**Authors:** Igor Nesteruk

**Affiliations:** Institute of Hydromechanics, National Academy of Sciences of Ukraine, Kyiv, Ukraine

**Keywords:** COVID-19 pandemic, COVID-19 mortality, COVID-19 epidemic dynamics in USA, China, India, France, Germany, Brazil, South Korea, Japan, Italy, UK, mathematical modeling of infection diseases, endemic characteristics of SARS-CoV-2, statistical methods

## Abstract

Current WHO reports claim a decline in COVID-19 testing. Many countries are reporting no new infections. In particular, USA, China and Japan have registered no cases and COVID-19 related deaths since May 15, 2023. To discuss consequences of ignoring SARS-CoV-2 infection, we compare endemic characteristics of the disease in 2023 with ones estimated before using 2022 datasets. The accumulated numbers of cases and deaths reported to WHO by 10 most infected countries and global figures were used to calculate the average daily numbers of cases and deaths per capita (*DCC* and *DDC*) and case fatality rates (*CFR*) for two periods in 2023. The average values of daily deaths per million still vary between 0.12 and 0.41. It means that annual global number of COVID-19 related deaths is still approximately twice higher than the seasonal influenza mortality. Increase of *CFR* values in 2023 show that SARS-CoV-2 infection is still dangerous despite of increasing the vaccination level. Very low *CFR* figures in South Korea and very high ones in the UK 4 need further investigations.

## Introduction

The high numbers of circulating SARS-CoV-2 variants [1-3] and re-infected persons [4-6], the lack of decreasing trends in the global numbers of deaths [7], and the expected very long duration of the Omicron wave [8] caused an assumption that the pathogen will circulate forever, which was supported by a mathematical model, [9]. Some endemic characteristics of the SARS-CoV-2 disease were estimated in [9] with the use of global accumulated numbers of COVID-19 cases *V* and deaths *D*, registered in [10] from January 1, 2022 to December 6, 2022. In particular, that the global numbers of new daily cases will range between 300 thousand and one million, daily deaths – between one and 3.3 thousand. Dividing these values by the world population 8060.5 million [11], we can calculate lower and upper limits per capita: for global average number of daily cases per million *DCC*^*\**^ = 37.2 and *DCC*^*\*\**^ = 124.1, and for the global average number of COVID-19 related deaths per million *DDC*^*\**^ = 0.124 and *DDC*^*\*\**^=0.409. The case fatality risk *CFR* (the ratio of number of deaths to the number of cases) was estimated in [9] for the figures listed in [10]. In particular, the global value *CFR*^*\**^=0.0033 for the period from January 1, 2022 till December 6, 2022.

In this study, we will try to check the estimations of [9] with the use of data reported by WHO in 2023, [7]. Unfortunately, currently “reported cases do not accurately represent infection rates due to the reduction in testing and reporting globally”, [12]. In particular, in the period from 31 July to 27 August 2023 only 39% of countries reported at least one case to WHO, [12]. Between 10 countries with the highest figures of accumulated COVID-19 cases three (USA, China and Japan) have stopped to update data after May 15, 2023 (see Table 1). Nevertheless, we will try to analyze recent trends for *DCC, DDC* and *CFR*, to compare them with *DCC*^*\**^, *DCC*^*\*\**^, *DDC*^*\**^, *DDC*^*\*\**^, and *CFR*^*\**^ and to answer the question: can we start to ignore the SARS-CoV-2 disease?

**Table 1.**
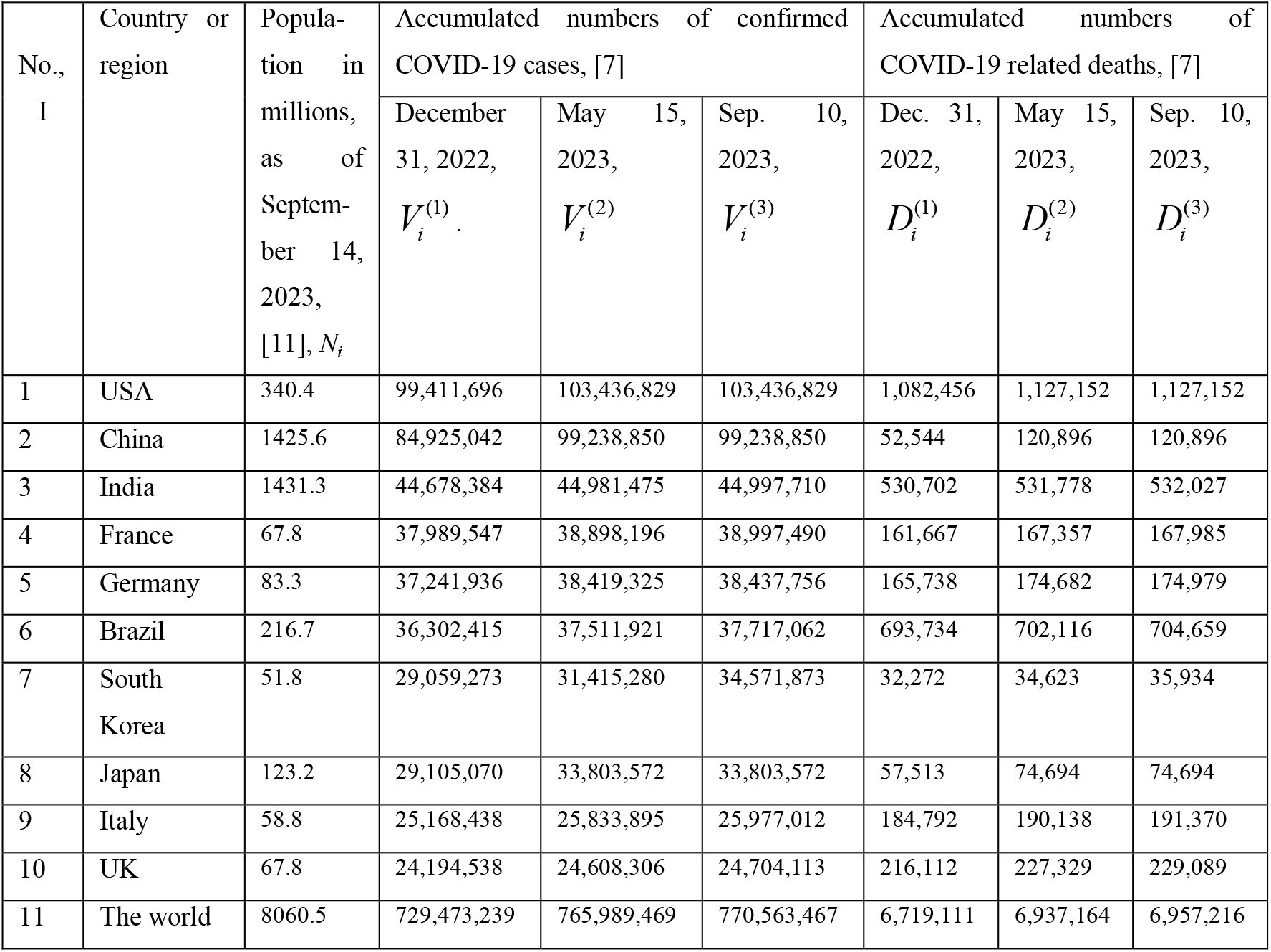
Accumulated numbers the COVID-19 cases and deaths and population volumes in 2023.

## Materials and Methods

We will use the accumulated numbers of laboratory-confirmed COVID-19 cases *V*_*i*_ and deaths *D*_*i*_ for 10 countries with the highest figures and the world (*i* =1, 2,.., 11) listed by WHO, [7] (version of file updated on September 14, 2023). Chinese figures include Mainland China, Taiwan, Hong Kong and Macao. 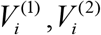 and 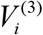 correspond to the numbers of cases accumulated on December 31, 2022; May 15, 2023 and September 10, 2023, respectively. 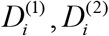 and 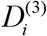 correspond to the accumulated numbers of deaths (see Table 1). The populations *N*_*i*_ of regions (as of September 14, 2023, [11]) are also listed in Table 1.

To calculate the averaged daily characteristics *DCC* and *DDC* we will use two periods: January 1 - September 10, 2023 (*T*_*1*_ =253 days) and May 16 - September 10, 2023, (*T*_*2*_ =119 days) and simple formulas:

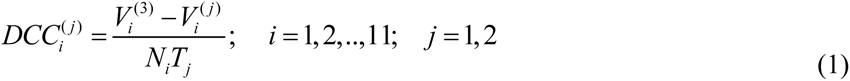

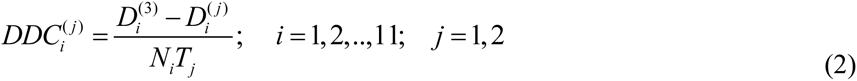

The CFR values corresponding to the same periods can be calculated as follows:

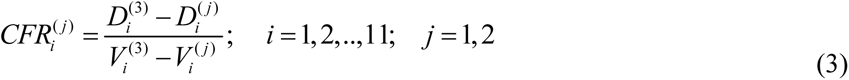

## Results and discussion

The results of calculations with the use of Eqs. (1)-(3) are presented in Table 2 and Fig. 1. Blue, black and red markers correspond to the values of *DCC, DDC* and *CFR*, respectively. The characteristics, corresponding to the long period (January 1 - September 10, 2023; *T*_*1*_ =253 days) are shown by “circles”; to the short period (May 16 1 - September 10, 2023; *T*_*2*_ =119 days) – by “crosses”. For comparison, the values estimated in [9] with the use of figures registered in 2022 are represented by dashed lines: blue for *DCC*^*\**^ and *DCC*^*\*\**^; black for *DDC*^*\**^ and *DDC*^*\*\**^; red for the case fatality risk *CFR*^*\**^=0.0033. Please pay attention on zero values of 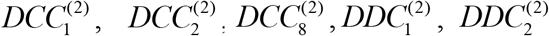, and 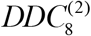 (not shown in Fig. 1) corresponding to the short period in USA, China and Japan, when the numbers of cases where not updated. Zero values of denominator in Eq. (3) does not allow calculating 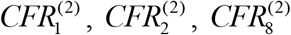.

**Table 2.**
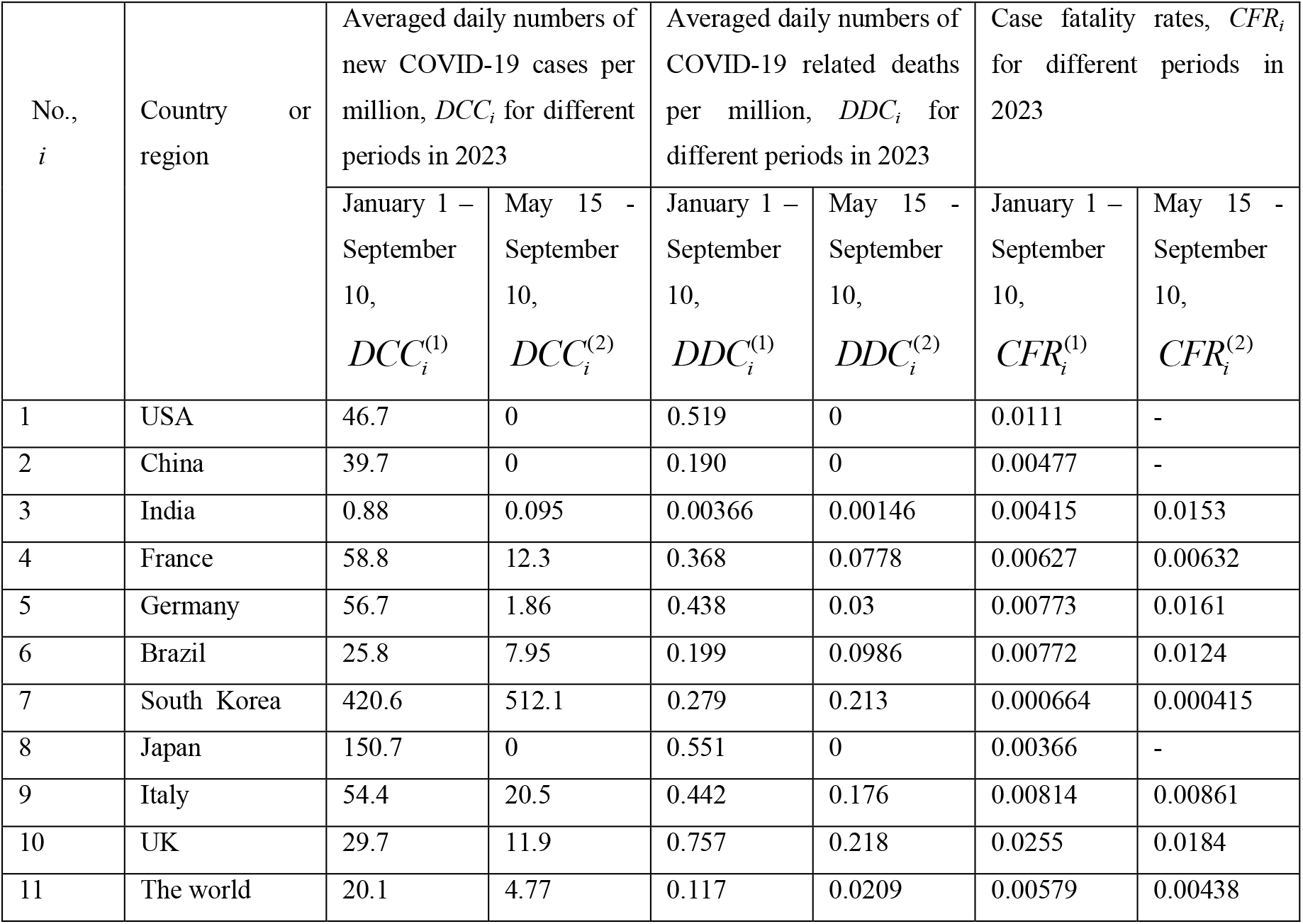
Results of calculations the averaged daily numbers of cases and deaths and case fatality rates for two different periods in 2023.

**Fig. 1.**
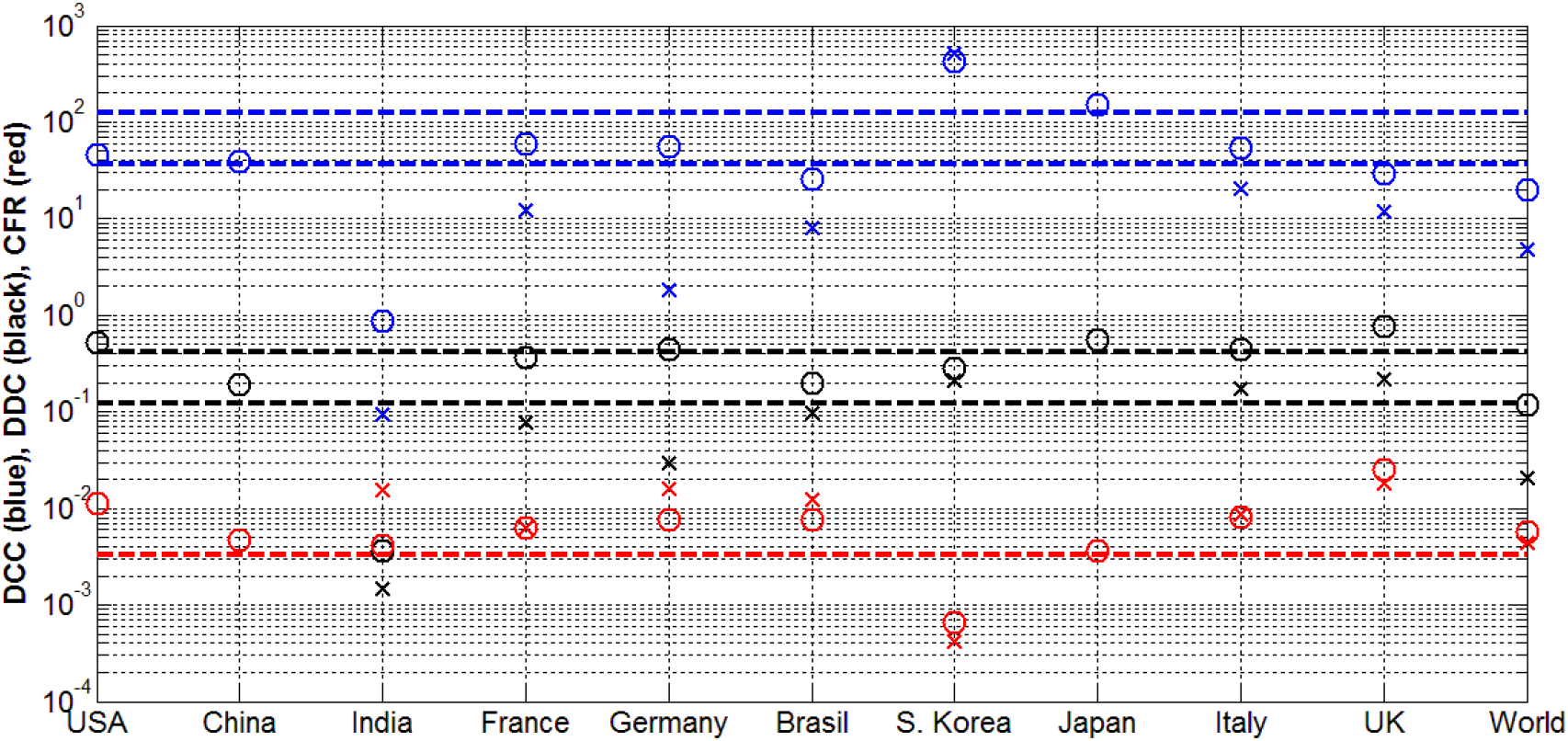
Results of calculations (markers) the averaged daily numbers of cases *DCC* (blue) and deaths *DDC* (black) and case fatality rates *CFR* (red) for two different periods in 2023 and comparison with the values estimated in [9] (lines). The characteristics, corresponding to the long period (January 1 - September 10, 2023; *T*_*1*_ =253 days) are shown by “circles”; to the short period (May 16 - September 10, 2023; *T*_*2*_ =119 days) – by “crosses”.

It can be seen that average values of new daily cases per capita corresponding to long period (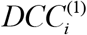, blue “circles”) are mostly close to the estimation *DCC*^*\**^ =37.2 (the lower blue line). The value 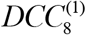 corresponding to Japan is slightly higher than 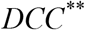 =124.1. In 2023, South Korea has registered much more cases than the global averaged value in 2022 (compare the corresponding blue “circle” and “cross” with the upper blue line).

The averaged daily numbers of cases per capita corresponding to the short period are zero for USA, China and Japan (not shown in Fig. 1) or much lower than the estimation *DCC*^*\**^ =37.2 (compare blue “crosses” and the lower blue line). There are two possible reasons of this result. As mentioned below, more and more countries reduce testing and stop updating the COVID-19 statistics [12]. On the other hand, the short period corresponds to the summer time in the most infected countries. Probably, we will see some increase in the end of 2023.

Very low numbers of cases and deaths registered in India is exclusion (compare the corresponding blue and black “circles” and “crosses” with the blue and black lines). Probably, this fact is connected with a low testing level. It was shown in [13] that insufficient testing does not allow detecting a lot of COVID-19 cases and deaths in poor countries (e.g., in Africa). Similar trend is visible in other countries and worldwide. In particular, reducing in testing after May 15, 2023 caused much lower values of 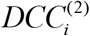 and 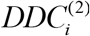 than 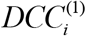 and 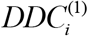, respectively (compare blue and black “crosses” and “circles”, South Korea is only one exception).

It can be seen that average values of daily deaths per capita corresponding to the long period (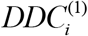, black “circles”) are mostly located between to the limits *DDC*^*\**^ and *DCC*^*\*\**^ (black lines). It means that these estimations (obtained in [9] with the use of 2022 datasets) still correspond to the real number of deaths. Multiplying the limits 1 to 3.3 thousand for global average daily deaths (calculated in [9]) by 365, the expected number of annual global COVID-19 related deaths varies between 365 thousand and 1.2 million. Unfortunately, these figures exceed the seasonal influenza mortality (between 294 and 518 thousand annual deaths in the period from 2002 to 2011, [14]).

Zero or much lower numbers of deaths during the short period (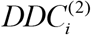, black “crosses”) reflect the decrease of registered cases due to reduce in testing. It does not mean that the number of deaths was much lower in the summer of 2023, but illustrates the fact that many of them were not revealed. Probably, it will be possible to detect an increase in this hidden mortality (registered without direct connection to the SARS-CoV-2 disease).

Much higher case fatality rates in 2023 (compare red “crosses” and “circles” with red line corresponding to 2022; South Korea is only one exception) show that SARS-CoV-2 infection is still dangerous despite of increasing vaccination level (more than 13.5 billions doses have been administered as of 31 August 2023, [12]). Higher *CFR* values can also be a result lower testing levels. Reducing the denominator in formula (3) can be much larger than nominator, since severe COVID-19 cases (leading to deaths) are easier to be detected. In any case, there are many issues to be clarified. In particular, why *CFR* values in South Korea are approximately 10 times higher and in the UK 4 times higher than the global figures.

## Conclusions

The endemic characteristics of SARS-CoV-2 infection in 2023 agree with ones estimated in [9] with the use of 2022 datasets, but lower testing and reporting levels have to be taken into account. In particular, the average values of daily deaths per million still vary between 0.12 and 0.41. It means that annual global number of COVID-19 related deaths is still approximately twice higher than the seasonal influenza mortality.

Increase of the case fatality rates *CFR* in 2023 show that SARS-CoV-2 infection is still dangerous despite of increasing the vaccination level. Very low *CFR* values in South Korea and very high figures in the UK 4 need further investigations.

## Clarification point

No humans or human data was used during this study

## Data availability

All data generated or analysed during this study are included in this text.

## Acknowledgement

The author is grateful to Oleksii Rodionov for his help in providing very useful information.

